# Initial stage of the COVID-19 virus infection process in human population

**DOI:** 10.1101/2020.04.13.20063701

**Authors:** S.A. Trigger

**Affiliations:** Joint Institute for High Temperatures, Russian Academy of Sciences, 13/19, Izhorskaia Str., Moscow 125412, Russia; Institut für Physik, Humboldt-Universität zu Berlin, Newtonstraße 15, D-12489 Berlin, Germany

## Abstract

The simple approximation for the first stages of the infection spread is considered. The specific feature of the COVID-19 characterized by its long latent period is taken into account. Exponential increase of numbers of infected people is determined by the half period of the maximal latent time for the COVID-19. The averaging over latent period leads to additional increase of the infected numbers.

**PACS number(s):** 02.50.-r, 05.60.-k, 82.39.-k, 87.19.Xx

## I. INTRODUCTION

The new virus named SARS-CoV-2 (causing COVID-19 disease) appeared first in China in December 2019 and to this moment (17 March 2020, 11:29 GMT) led to 186665 confirmed cases of disease and 7467 deaths in the World. The percent of the deathes is unusually high, especially in the group of old people, in comparison with the other flu infection associated with other viruses (see the website https://www.worldometers.info/coronavirus/coronavirus-cases/). 11 March 2020 this disease is recognized by the World Health Organization (WHO) as a pandemic. We consider the simplest model of the COVID-19 spread which takes into account a long latent period of this disease.

There are various scenario and models for the description of the epidemic situation. The most models are concentrated on the spontaneous development of the infection spread to describe all stages of the process. An example is, e.g., the susceptible-infected-susceptible (SIS) - one of the basic themes in the mathematical epidemiology [1,2]. These models describe the balance between the susceptible and infected individuals in population under the various conditions of infection transfer [3]. Kinetic approach can be applied on the basis of works [4], [5]. However, the situation with the COVID-19 has the specific features which should be taken into account.

There are essential efforts to confront the epidemic, to localize its spread. In general we can characterize its development as a non-spontaneous epidemic process. It is especially important also to slow down the propagation velocity to enable medicine to provide treatment for patients. Under these conditions the investigation of the initial stage of infection is needed. The long latent period of the COVID-19 is the feature which is considered in this paper.

## II. THE INFECTION DEVELOPMENT

If the number of dangerous (at distance less *s* ≃ 2 meters) contacts per day equals *N*_*c*_ for one infected person and *k* is the coefficient of infection transfer (the ratio of the number of people accepting virus to the number of dangerous contacts *N*_*c*_) we find that the average number of infected persons in population after *l*_0_ days can be estimated as the half of the “maximal latent time” 2*l*_0_ days for the COVID-19.

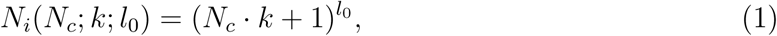

where *l*_0_ is the average number of days between obtaining the virus and appearance of the illness after which persons have to be isolated. Note, the the case of *N*_*c*_ · *k* ≫ 1 has been considered in [6].

As an example, let us consider *N*_*c*_ = 50, *k* = 0.1 and *l*_0_ = 7 (based on the half of the maximal latent period equals of 14 days). As easily seen, in this example *N*_*i*_(50; 0.1; 7) = 279936. If *N*_*c*_ = 25, *k* = 0, 1 and *l*_0_ = 7 we find *N*_*i*_(25; 0.1; 7) = 6434. The value of the parameter *k* is unknown. It can be roughly estimated but practically cannot be changed.

The average value *l* = *l*_0_ is well known, but of course, there is some distribution on *l*, which is not included in the simplest model under consideration. In Fig.1 the dependence of the infected cases *N*_*i*_ on the number of dangerous contacts *N*_*c*_ per one infected person is shown in the typical interval 50 ≥ *N*_*c*_ ≥ 0. The various curves correspond to the coefficient of infection transfer (or probability of infection) equals *k* = 0.1, 0.08, 0.07.

**Figure 1:**
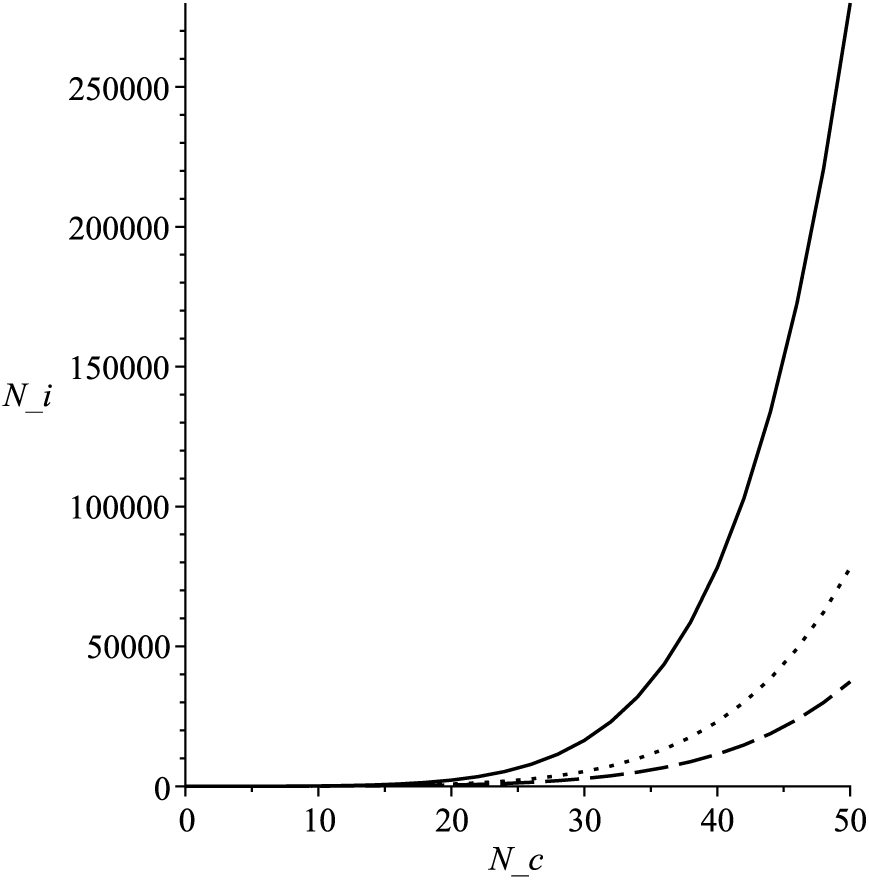
Infected cases *N*_*i*_ versus the dangerous contacts *N*_*c*_ for the different coefficient of infection transition *k*. The solid line corresponds to *k* = 0.1, the dot line to the value *k* = 0.08 and the dashed line *k* = 0.07 after *l*_0_ = 7 equals the half period of the maximal latent time.

To generalize the above consideration for the case of distribution on maximal latent period *l*, which always exists, we use the Gauss normalized distribution around *l* = *l*_0_ (the restriction for the maximal latent period is removed due to exponential decrease of the distribution)

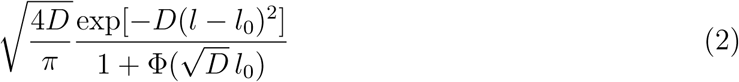

where Φ is the error function 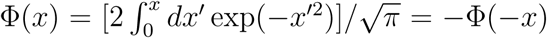. To find approximately the result for the average on *l* value of infected people ⟨*N*_*i*_(*N*_*c*_; *k*; *l*)⟩_*l*_ we can transfer to the continuous description on *l* and calculate the integral (*a* ≡ *N*_*c*_ · *k* + 1)

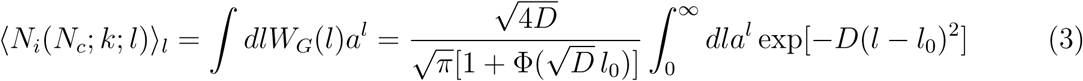

Using the stationary-phase method we arrive at the approximate result for the the integral in Eq. (3)

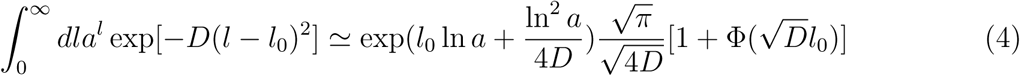

Therefore, for the averaged over the Gauss distribution *l* number of infected people in population is given by

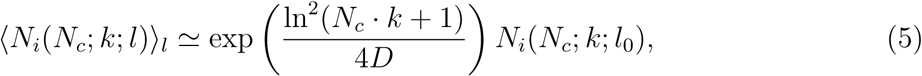

that is generalization of Eq. (1). For *D* → ∞ we arrive to the result of Eq (1). As is clear the distribution on *l* always leads to increase of the infected cases ⟨*N*_*i*_(*N*_*c*_; *k*; *l*)⟩_*l*_ in comparison with *N*_*i*_(*N*_*c*_; *k*; *l*_0_). The curves in Figure 2 illustrate the role of averaging over l with different values of the averaging parameter D=0.1 and D = 0.05 (shown, correspondingly, in solid and dashed lines) and their comparison with the non-averaged curve (dotted line - the function *N*_*i*_(*N*_*c*_; *k*; *l*_0_ = 7) with *N*_*c*_ = 20 ÷ 50 and *k* = 0.08. The difference between ⟨*N*_*i*_(*N*_*c*_; *k*; *l*)⟩_*l*_ and *N*_*i*_(*N*_*c*_; *k*; *l*_0_ = 7) increases with *N*_*c*_ increase and can be essential for *N*_*c*_ *>* 40 for the considered parameters.

**Figure 2:**
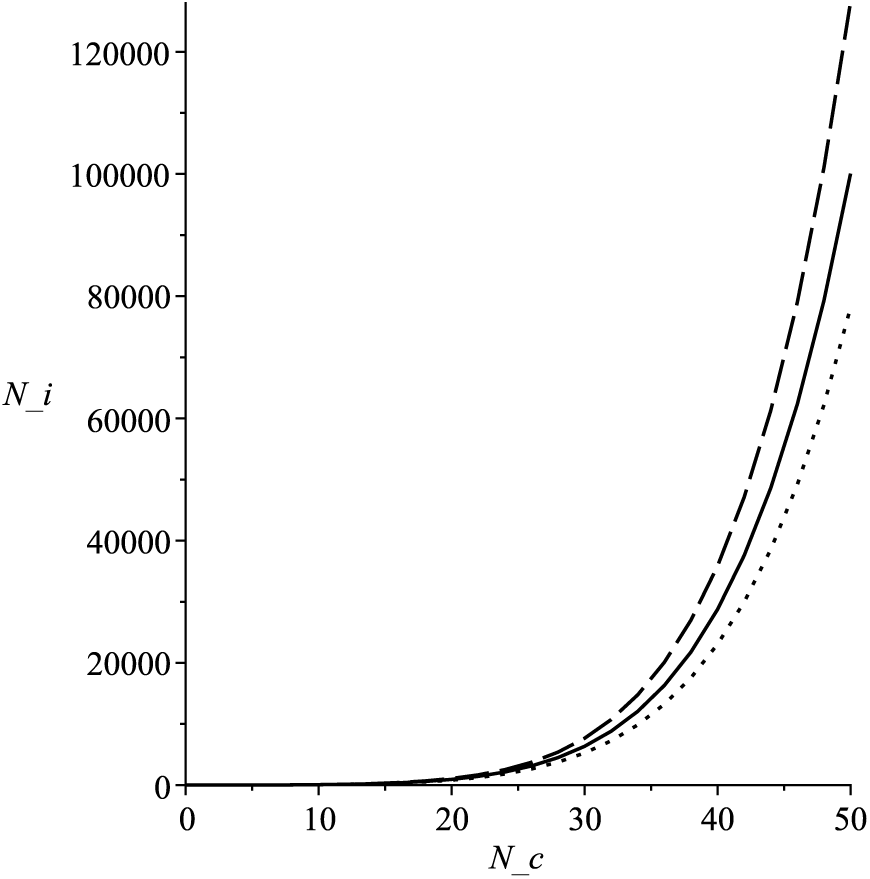
Averaged over *l* infected cases ⟨*N*_*i*_(*N*_*c*_; *k*; *l*)⟩_*l*_ versus the dangerous contacts *N*_*c*_ for the coefficient of infection transit *k* = 0.08 and the parameter *D* = 0.1 (solid line) and *D* = 0.05 (dashed) in comparison with the curve *N*_*i*_(*N*_*c*_; *k* = 0.08; *l*_0_ = 7) (the dotted line which coincides with the same on Fig. 1)

## III. CONCLUSIONS

The typical for the virus SARS-CoV-2 long maximal latent period 2*l* leads to the fast exponential increase of the average infected people with the exponent *l*. The subsequent restriction of the infection spread is related with the fast and true quarantine actions, which transfer first the exponential increase to saturation, as it is in China at the moment and to further decrease.

## Data Availability

I aprove in this statement the availability of all data reffered in the manuscript

## Acknowledgment

Author is thankful to M.V. Fedorov and A.M. Ignatov for the useful discussions.

